# Priorities for AI Education: Clinicians’ Perspectives

**DOI:** 10.1101/2025.07.29.25330662

**Authors:** Mark Jeffrey, Eric Auyoung, Denys Pak

**Affiliations:** Digital Health Fellow, NHS England East of England; Department of Anaesthesia and Critical Care, Peterborough City Hospital, UK; Department of Radiology, The Princess Alexandra Hospital NHS Trust, UK; Postgraduate Associate Dean, NHS England East of England

**Keywords:** [artificial intelligence, Decision Support Systems Clinical, Information Literacy, Health Personnel]

## Abstract

**Objective:** Educating clinicians about Artificial Intelligence (AI) is an urgent need(1) as the UK General Medical Council (GMC) places liability with practitioners(2) and the EU AI Act with employers to provide appropriate training(3), but also because AI, like any tool, requires training to use safely. NHSE Capability Framework provides guidance(4), but frontline clinicians’ perspectives are unknown so we sought to identify their priorities.

**Methods and Analysis:** Using iterative interviews with residents, educators and experts we synthesised 10 contextualised AI-related problem statements. We surveyed residents and consultant-educators in the East of England, who rated their confidence and importance. Participants also ranked their preferred learning modality.

**Results:** We received 299 responses. Clinicians priorities, defined by high importance (I) and low confidence (C), were: ‘understanding liability implications’ (I: 40%; C: 1.82/5), ‘determining appropriate levels of confidence in AI algorithms’ (I: 36.5%; C: 1.98/5), and ‘mitigating security and privacy risks’ (I: 34%; C: 1.68). Confidence was low (mean 20, range 10-50), with no significant difference between educators and residents. Residents preferred integration of training into regional teaching, while consultant-educators favoured webinars.

**Conclusion:** Our findings show that clinicians prioritise practical concerns, such as liability and determining confidence in algorithmic outputs. In contrast, critical appraisal and explaining AI to patients were deprioritised, despite their relevance to clinical safety. This study enhances the NHSE Capability Framework by contextualizing AI-related capabilities for clinicians as users and identifying priorities with which to develop scalable training.

**Key Messages:** *What is already know on this topic:* While clinicians face legal accountability for their use of AI in healthcare(2,3,5), there remains no standardised educational pathway to support them in acquiring the necessary skills. Although expert-informed capability frameworks exist(6), they are necessarily broad and lack operational clarity for day-to-day clinical roles.

*What this study adds:* This study translates 31 AI-related capabilities from the NHSE DART-Ed Capability Framework(6) into 10 concise AI learning needs for clinicians of the user archetype through iterative interviews with residents, educators and AI experts. A regional survey with 299 responses from residents and educators highlights practical concerns such as liability and determining appropriate confidence in AI algorithms as learners’ priorities, whilst critical appraisal and explaining AI to patients were deprioritised despite their relevance to clinical safety.

*How this study might affect research, practice or policy:* The educational priorities of clinicians as users of AI identified in this study provides engaging, curriculum-ready content mapped to the ‘user’ archetype of the DART-Ed framework, which can be adapted to role and task-specific educational activities.

## Introduction

Despite the growing use of AI by clinicians in real-world care settings, there remains no consistent, curriculum-aligned framework to ensure frontline staff are adequately trained, legally protected, or ethically equipped to use these tools safely. Key NHS strategy documents, including the 2019 NHS Long Term Plan(7) and the subsequent 2019 Topol Review(8), emphasise digital transformation and Artificial Intelligence as crucial to improving patient care, but acknowledge significant implementation barriers.

Education and training for frontline staff are repeatedly highlighted as crucial to realising these improvements. This has been further emphasised by the EU AI Act giving employers responsibility to ensure staff using AI systems are appropriately trained(3). The General Medical Council (GMC) emphasises that clinicians retain liability for AI-informed decisions, stressing the need to practice within their competence and maintain up-to-date knowledge(2).

In response, NHS England (NHSE) Department of Workforce Training, Education and Development (formerly Higher Education England) set up the Digital, Artificial Intelligence and Robotics Technologies in Education (DART-Ed) Programme and commissioned a learning needs analysis and formulation of a capabilities framework to support the development of digital skills and literacy for healthcare workers(6). While the framework outlines high-level capabilities, these have not yet been translated into specific, role-aligned learning objectives that can guide curriculum development or policy implementation.

Recent advances in AI have led to the widespread use of general-purpose Large Language Models (LLMs), like ChatGPT, resulting in rapid uptake that has outpaced clinician training in its use. Indeed, a recent clinician survey reported that over half of respondents routinely use LLMs in clinical practice despite receiving no formal guidance on data governance, hallucinations, liability, or appropriate educational supervision for trainees(9).

Given our responsibility to deliver training to Resident Doctors and Educators in the East of England (EoE), Health Innovation and NHSE EoE sought to better understand the Learners’ perspectives and Educator’s preparedness to deliver training, aligned specifically with the frontline clinician or ‘User’ archetype defined within the DART-Ed Framework. Therefore, our aim was to identify the most relevant, high-impact problem statements that map directly to the framework.

After formulating concise problem statements relevant to Clinicians as “Users” of AI, we surveyed Residents and Consultant Educators in the EoE, to gauge their baseline confidence and perceived importance. By combining perceived importance and self-reported confidence, we aimed to prioritise educational content that is both high-stakes and under-addressed, thereby laying the foundation for a reproducible, domain-specific AI curriculum for clinicians.

## Materials and Methods

The DART-Ed AI and Digital Health Capabilities Framework was reviewed, filtering capability statements aimed at the “User” archetype and which related to Clinical AI. This produced a list of 31 capability statements from the 198 in the whole framework. In order to contextualize these problem statements for resident doctors in the East of England, and to facilitate feedback from learners and educators we developed 10 concise Problem statements that linked to the relevant capabilities (Appendix 1). We conducted interviews with 5 residents, 3 senior educators and 7 clinical AI experts to iteratively gather feedback on these problem statements. Problem statements were reviewed and edited based on feedback between interviews.

A cross-sectional survey of Resident Doctors and Consultant Educators was conducted in the East of England. The survey consisted of 9 questions (Appendix 2). We asked participants to self-rate their confidence in each problem on a 1-5 likert scale and to choose up to 3 problem statements they felt were most important to their clinical practice, rank their preferred teaching modality, as well as their training grade, speciality and educational role. White space responses will be thematically analysed separately. Invitations were sent through NHSE EoE mailing lists in a single stage sample. NHS email account verification was required to complete the survey. Questions were reviewed and edited by the authors but formal pre-testing was not performed. Responses were collected between December 2024 to February 2025.

Responses to training grade and educator tier questions were dichotomised into consultant educators and residents for the primary comparison of Resident versus Educator. All respondents who indicated they were a Consultant or General Practitioner (GP) AND Educator Tier ≥ 2(10) (Clinical Supervisor or higher) were included in the “Educator” group, whilst all non-consultant/GP training grade responses were included in the “Resident” group.

### Statistical Methods

The primary comparison was of Educator vs Resident. Secondary analyses were performed by training grade, educator tier and specialty. Missing responses were excluded and no weighting of items or propensity score adjustments were made. Student’s T-test was used to compare importance votes and confidence ratings. Bonferroni correction was applied to adjust P-values for multiple comparisons. The threshold for statistical significance was set at adjusted P < 0.05. Statistical analysis was done using R Statistical Software v4.4.3(11) with tidyverse v2.0.0(12) and rstatix v0.7.2(13) packages.

### Ethics and Data Management

The MRC and NHS Health Research Authority decision tool was consulted indicating NHS research and ethics committee review was not required for this project. An IRB equivalent of Health Education England, East of England waived ethical approval for this work. No patients were involved in the study. A consent statement for research use of anonymised responses was included in the survey introduction. Responses were collected using an NHS.net Microsoft Forms account, with access to results restricted to the authors. NHS.net email validation was required to complete the survey, but responses were collected anonymously. Identifiable demographic data was intentionally minimised to limit the risk of de-anonymization and limited to clinical speciality, training grade and educator tier(14). Free text responses were manually screened and any identifiable information such as email addresses were censored.

### Patient and Public Involvement

As we are investigating resident doctors and their educators perspectives we did not have direct public or patient involvement in this study. We build on a framework(6) developed with significant academic and industry involvement.

## Results

### Problem Statements

After review of the DART-Ed framework and iterative feedback from 15 interviews with Residents, Educators and AI experts, the final 10 problem statements (in no particular order) where:

1. I am able to determine an appropriate level of confidence in a decision or result from an AI algorithm
2. I am able to write effective prompts for Generative AI and recognise AI generated content
3. I am aware of realistic use cases for AI in healthcare
4. I can critically appraise papers reporting AI interventions in my speciality
5. I can explain how AI in healthcare is regulated
6. I can explain to patients how AI works
7. I can identify sources of bias in AI algorithms and potential mitigation measures
8. I can mitigate security and privacy risks posed by AI
9. I know common ways that AI can fail
10. I know the liability implications of using AI in my practice

Together these map on a many-to-one basis to the 31 relevant capabilities in the DART-Ed framework (Appendix 1).

### Survey

#### Response Rate and Demographics

We had 317 responses to the survey, which was shared via NHSE EoE mailing lists to ∼7000 resident doctors and ∼5000 consultant educators in the region (overall response rate ∼2.4%). We received 151 responses from consultant educators (∼3% response rate) and 145 from residents (∼2.1% response rate) with 21 (0.1%) not reporting their training grade (Figure 1). Training grades included 14 responses from Foundation Doctors, 75 from ST/CT1-3, 43 from ST4-6, 11 from ST7+ and 153 from Consultants (2 responding consultants reported no educational role). The Educator Tier included 91 non-educators, 53 Trainees as educators, 20 Clinical Supervisors, 96 Educational Supervisors, 51 College Tutors or Training Programme Directors and 6 Heads of school or Associate/Deputy Deans with no missing responses.

**Figure 1:**
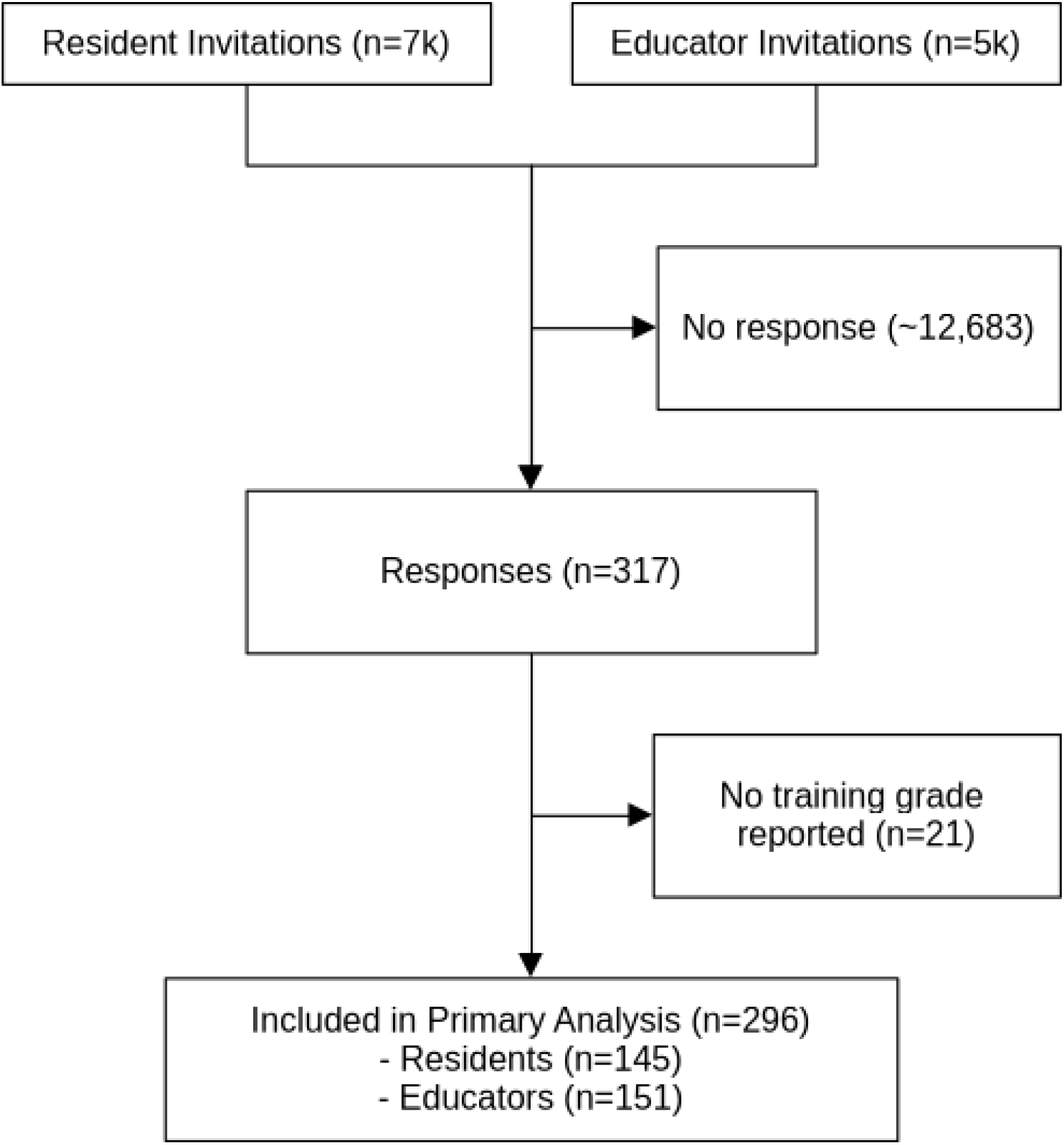
Survey consort diagram. We received responses from 33 different specialities (Figure 2). The largest groups were Internal Medicine (62 [19.6%]), General Practice (48 [15.1%]) and Surgery (31 [9.8%]). Numbers reporting Foundation Programme as a speciality showed a discrepancy from those reporting Foundation Programme as their training grade (17 vs 14). Speciality was not reported by 2 (0.6%) respondents, with 290 (91.5%) reporting a single speciality, 21 (6.6%) reporting 2 specialities and 4 (1.3%) reporting 3 specialities.

**Figure 2:**
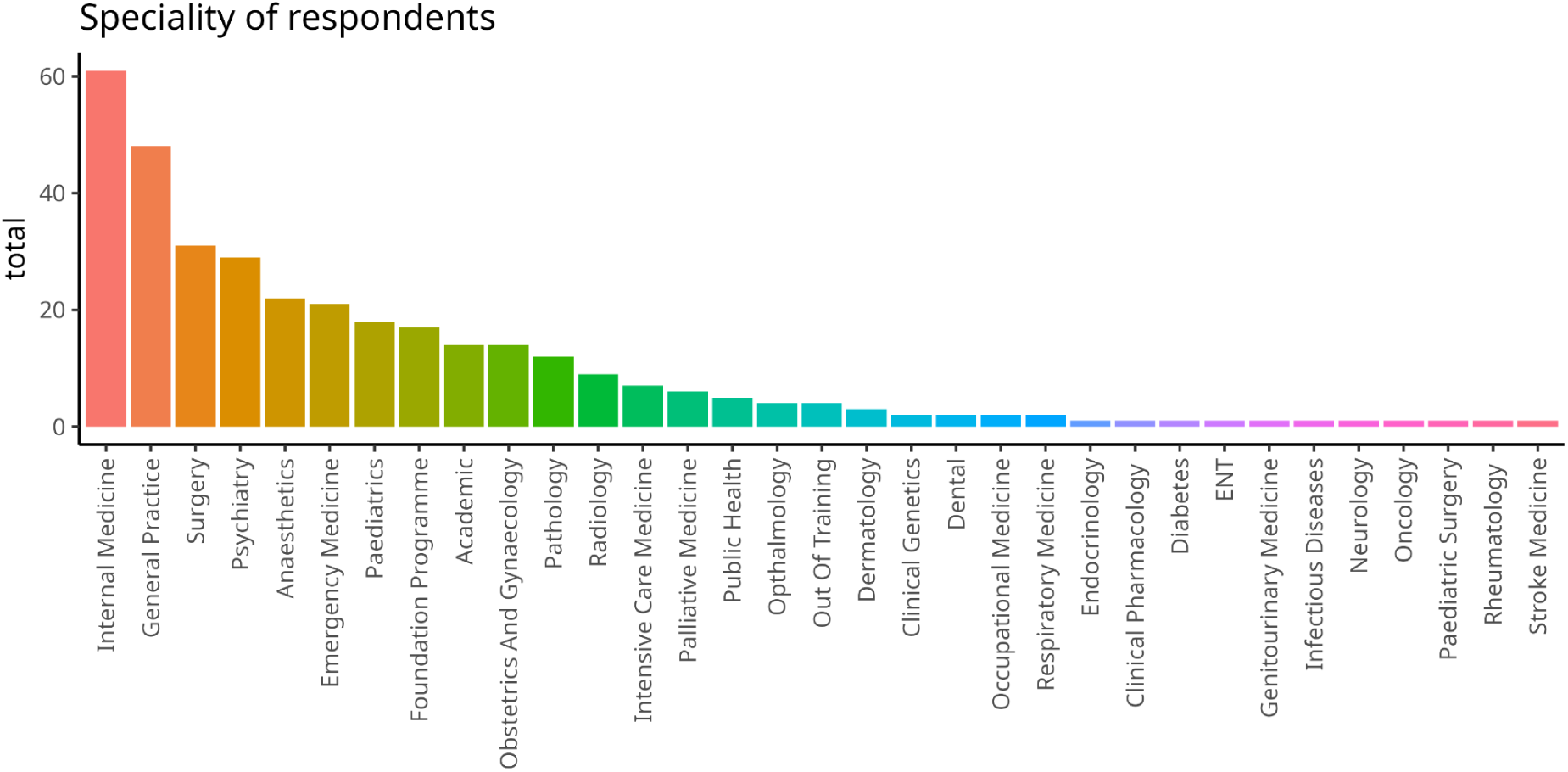
Bar chart showing speciality reported by each respondent ranked by number of responses. All respondents rated their confidence in each statement. For importance votes, 9 (2.8%) respondents did not vote for any of the statements, 4 (1.3%) voted for one, 5 (1.6%) voted for 2 and 299 (94.3%) voted for the maximum of three. Regarding teaching format preferences 48 (15%) did not respond.

#### Confidence Ratings

Overall confidence ratings were low (Table 1, Figure 3), but there was significant heterogeneity between respondents. Mean total confidence score was 20.2/50 (95% CI: 19.2-21.3). Highest mean confidence was reported for “I am aware of realistic use cases for AI in healthcare” 2.50/5 (95% CI: 2.36-2.64), “I know common ways that AI can fail” 2.38/5 (95% CI: 2.25-2.51) and “I can explain to patients how AI works” 2.29/5 (95% CI: 2.15-2.42). Lowest mean confidence was reported for “I can explain how AI in healthcare is regulated” 1.55/5 (95% CI: 1.45-1.65), “I can mitigate security and privacy risks posted by AI” 1.68/5 (95% CI: 1.57-1.78) and “I know the liability implications of using AI in my practice” 1.81/5 (95% CI: 170.-1.94). There were no statistically significant differences in confidence comparing residents to educators, by training grade, educator tier or speciality.

**Table 1:** Confidence ratings on a likert scale (range 1-5) by problem statement ranked in descending order. “Total” is the sum of all confidence ratings reported by an individual (range 10-50).

| **Confidence Ratings** | **N** | **Mean** | **95% CI** | **Lower** | **Upper** |
| --- | --- | --- | --- | --- | --- |
| total | 296 | 20.226 | 1.031 | 19.195 | 21.257 |
| I am aware of realistic use cases for AI in healthcare | 296 | 2.503 | 0.139 | 2.364 | 2.642 |
| I know common ways that AI can fail | 296 | 2.378 | 0.133 | 2.245 | 2.511 |
| I can explain to patients how AI works | 296 | 2.287 | 0.134 | 2.153 | 2.421 |
| I am able to write effective prompts for Generative AI and recognise AI generated content | 296 | 2.152 | 0.141 | 2.011 | 2.293 |
| I can critically appraise papers reporting AI interventions in my speciality | 296 | 2.007 | 0.134 | 1.873 | 2.141 |
| I am able to determine an appropriate level of confidence in a decision or result from an AI algorithm | 296 | 1.98 | 0.126 | 1.854 | 2.106 |
| I can identify sources of bias in AI algorithms and potential mitigation measures | 296 | 1.875 | 0.12 | 1.755 | 1.995 |
| I know the liability implications of using AI in my practice | 296 | 1.818 | 0.122 | 1.696 | 1.94 |
| I can mitigate security and privacy risks posed by AI | 296 | 1.676 | 0.108 | 1.568 | 1.784 |
| I can explain how AI in healthcare is regulated | 296 | 1.551 | 0.1 | 1.451 | 1.651 |

**Figure 3.**
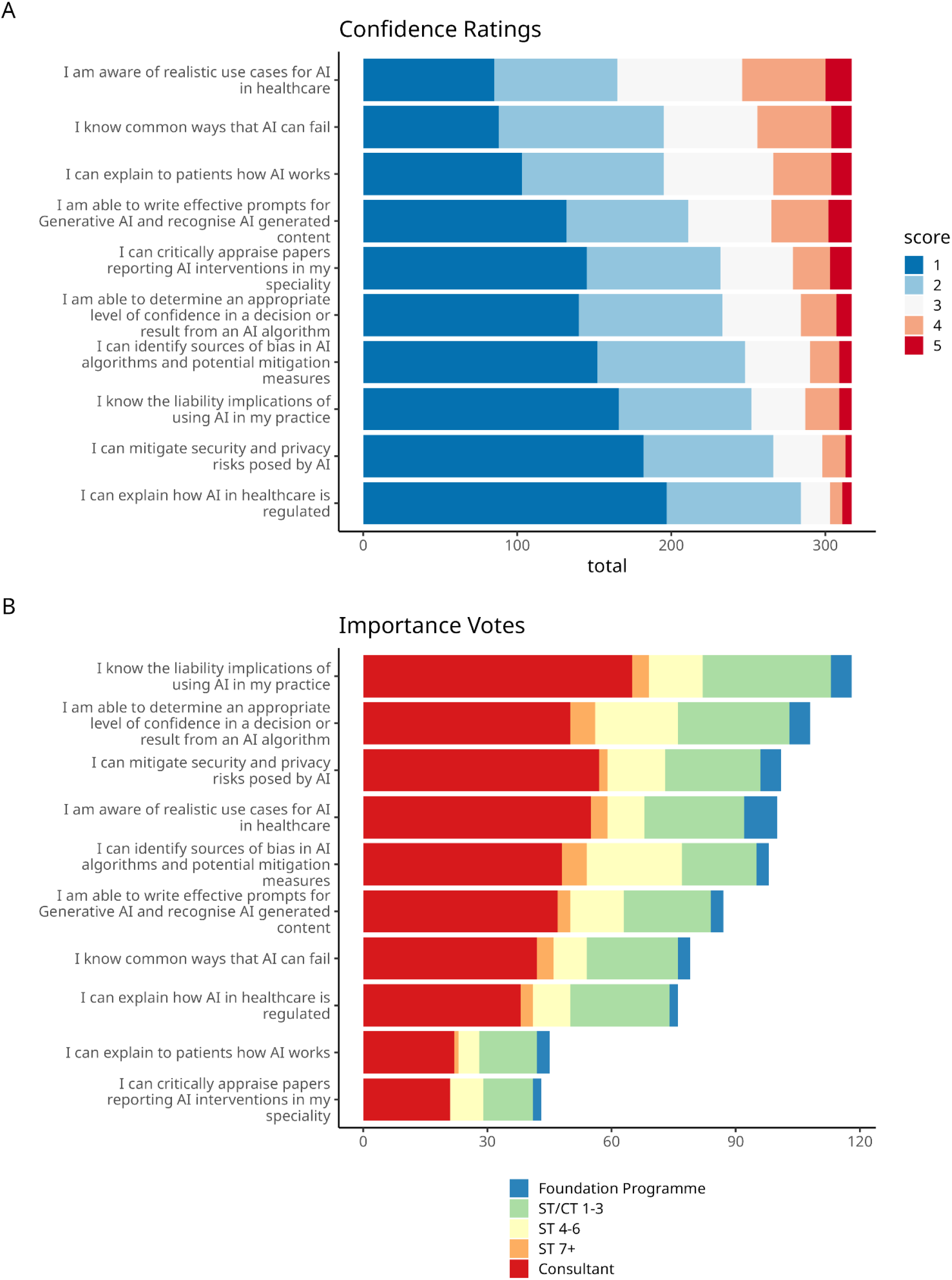
(A) Stacked bar chart of confidence rated on a 1-5 Likert scale from 1=“Not at all confident” to 5=“Completely Confident” for each problem statement ranked by mean confidence. (B) Stacked bar chart of total number of votes for each problem statement ranked by total and coloured by Training Grade.

#### Importance Votes

The three problem statements rated most important where “I know the liability implications of using AI in my practice” 118/296 (39.9%, 95% CI: 34%-46%), “I can determine an appropriate level of confidence in a decision or result form an AI algorithm” 108/296 (36.5%, 95% CI: 31%-42%) and “I can mitigate security and privacy risks posed by AI” 101/296 (34.1%, 95% CI: 29%-40%). Although 95% confidence intervals overlap for top 6 problems by importance voting (Table 2). Two problem statements were rated significantly less important than all others: “I can explain to patients how AI works” 45/296 (15.2%, 95% CI: 11%-19%), “I can critically appraise papers reporting AI interventions in my speciality” 43/296 (14.5%, 95% CI: 11%-19%) and “I can explain how AI in healthcare is regulated” 76/296 (26%, 95% CI: 21%-31%) (Figures 3-4).

**Figure 4.**
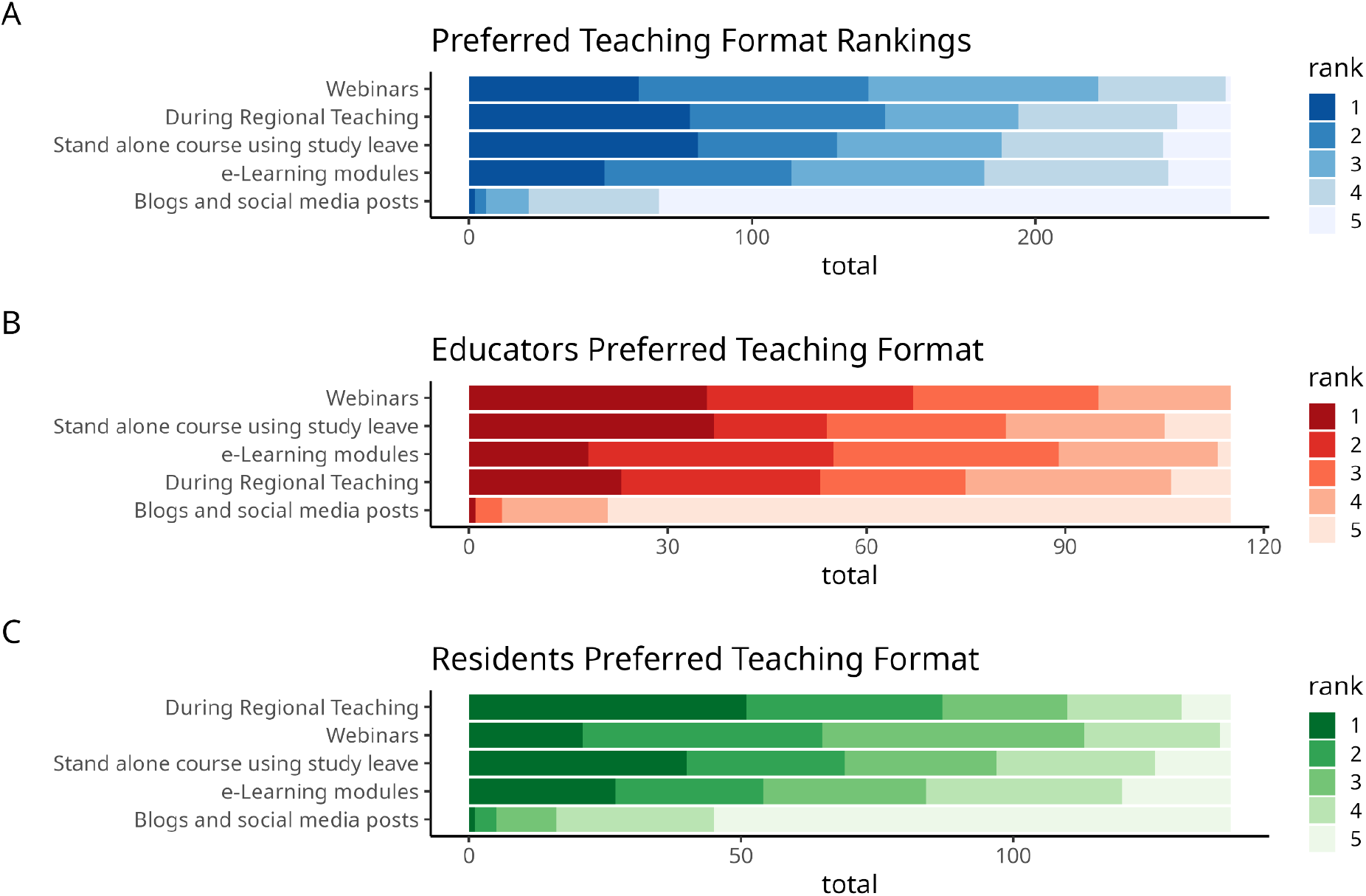
Teaching methods were ranked by each respondent from 1-5. (A) Stacked bar chart showing total number of respondents assigning each rank, (B) responses from educators, (C) responses from residents.

**Table 2:** Importance votes by problem statement ranked in descending order. Total is the absolute number of votes given to a problem statement. Mean shows the proportion of respondents who voted for a given problem.

| Importance Votes | Total | N | Mean | 95% CI | Lower | Upper |
| --- | --- | --- | --- | --- | --- | --- |
| I know the liability implications of using AI in my practice | 118 | 296 | 0.399 | 0.056 | 0.343 | 0.455 |
| I am able to determine an appropriate level of confidence in a decision or result from an AI algorithm | 108 | 296 | 0.365 | 0.055 | 0.31 | 0.42 |
| I can mitigate security and privacy risks posed by AI | 101 | 296 | 0.341 | 0.054 | 0.287 | 0.395 |
| I am aware of realistic use cases for AI in healthcare | 100 | 296 | 0.338 | 0.054 | 0.284 | 0.392 |
| I can identify sources of bias in AI algorithms and potential mitigation measures | 98 | 296 | 0.331 | 0.054 | 0.277 | 0.385 |
| I am able to write effective prompts for Generative AI and recognise AI generated content | 87 | 296 | 0.294 | 0.052 | 0.242 | 0.346 |
| I know common ways that AI can fail | 79 | 296 | 0.267 | 0.051 | 0.216 | 0.318 |
| I can explain how AI in healthcare is regulated | 76 | 296 | 0.257 | 0.05 | 0.207 | 0.307 |
| I can explain to patients how AI works | 45 | 296 | 0.152 | 0.041 | 0.111 | 0.193 |
| I can critically appraise papers reporting AI interventions in my speciality | 43 | 296 | 0.145 | 0.04 | 0.105 | 0.185 |

There were no statistically significant differences in importance votes between Educators and Residents. As a consequence of 0/11 ST7+ respondents voting for critical appraisal of papers, statistically significant differences after FDR correction were observed for this group compared to consultants and ST/CT1-3 groups. No other statistically significant differences between training grades, educator tier or speciality were identified for perceived importance of the problems.

#### Preferred Teaching Method

The teaching method with the highest mean rank were webinars (2.43), however regional teaching and stand alone courses were ranked 1st by more people (60, 78 and 81 respectively, Figure 6). There was a split between educators and residents on preference for regional teaching, which is not typically delivered to educators, with residents ranking regional teaching highest whilst educators ranked webinars, stand alone course and e-learning higher. Blogs and social media posts were consistently ranked lowest.

## Discussion

Understanding the perspectives of learners and educators is an important component in developing training for emerging technology. There is not yet a defined syllabus or consensus on what clinicians need to learn about AI in healthcare, despite the broad agreement about the importance of such education. The DART-Ed Artificial Intelligence and Digital Healthcare Technologies Framework(6) is necessarily broad, covering all categories of healthcare workers and engagement levels from front-line users to Department of Health strategists and computer scientists. This poses a challenge at the regional level when attempting to take the first steps in implementing training for specific groups and contexts.

To maximise engagement, we condensed the 31 relevant capabilities into a smaller number of problem statements. This also provided an opportunity to contextualize these for Clinicians of the “User” archetype. We selected this group as it is within the remit of Health Education England to train this group, and it seemed logical to develop widely applicable, basic resources for “users” before addressing higher engagement archetypes that are likely to have more specialised requirements.

Given the lack of definitive curricula or consensus on which components of AI are most important to teach, we felt identifying learners’ priorities was valuable. Delivering training aimed at topics learners identified as important and where confidence is low will maximise engagement. This engagement is the initial goal of early clinician education programmes in artificial intelligence.

We received a response rate higher than previous surveys carried out by Health Innovate East, suggesting that Residents and Educators are motivated to engage with training on AI in healthcare. However, a response rate of 317 out of ∼12000 doctors (∼7000 residents and ∼5000 educators, response rate ∼2.6%) may introduce selection bias with respondents with strong feelings about AI, both positive and negative, being more likely to complete the survey. Additionally the regional restriction on responses may impact generalizability of the results. We received balanced numbers of responses from Educators and Residents, facilitating comparison. As there are fewer Educators than Residents, this also indicates higher engagement with the survey from Educators (resident response rate 2.1%, educators 3%). There are a number of potential explanations for this including greater baseline interest in AI, educational projects or power asymmetry due to the request to complete coming from an Associate Dean rather than a Digital Health Fellow.

### Confidence

Confidence ratings were universally low, which is expected given the recency of AI developments and previously stressed importance of educating clinicians about AI. In fact the modal response was to give the minimum rating to all 10 problems. Despite this some respondents did report significant confidence levels, with 2 respondents giving maximum scores to all 10 problems. The problem with highest reported confidence (being aware of realistic use cases for AI in healthcare) also had the lowest cognitive demand based on Bloom’s Taxonomy(15), only requiring recall, and may explain the higher confidence scores. However, the problems with the highest cognitive load, requiring analysis and evaluation (such as critical appraisal, determining appropriate levels of confidence in AI algorithms decisions and identifying sources of bias in AI algorithms and potential mitigation measures) were not the lowest ranked.

The lowest confidence levels were reported for explaining how AI is regulated, mitigating security and privacy risks and knowing liability implications of using AI, which require understanding and application. It is possible that these governance related problems are perceived as more bureaucratic, technically dry or less clinically relevant and therefore respondents were less likely to have researched these areas, or felt less confident in non-clinical aspects. Alternatively, as governance of AI in healthcare is rapidly evolving and remains an unsolved problem it may be that these areas are harder to develop confidence in, especially with conflicting approaches to AI regulation emerging between the United States and European Union(16).

A key component of explaining to patients how AI works is communicating the privacy risks involved. UK General Data Protection Requirements (GDPR) requires that we disclose to patients who we are sharing their data with(17), and the recognition that the use of cloud-based AI services requires disclosure of sensitive health data to third parties should form an essential part of this patient explanation. This is required to fulfil both GMC ethical and legal standards for confidentiality(18) and UK GDPR consent to sharing, without which clinicians may find themselves liable to legal action. Given low confidence in liability and security and privacy risks, it appears there is poor awareness of what would constitute a satisfactory patient explanation about the use of AI systems in their healthcare, and the importance of consent to data sharing within this.

### Importance

Each respondent was able to vote for up to 3 problems as being most important for them. Therefore the proportion of votes indicates the percentage of respondents who included this problem in their top 3. Whilst this makes interpretation less clear than a single vote or ranking all 10, during survey testing and interview feedback many respondents felt all or most problems were important and found direct comparisons difficult. We therefore wanted to allow each respondent to give more than one response without forcing false dichotomies.

The interpretation of importance voting must be considered in the context of the survey participants - the majority of respondents are resident and educator clinicians who reported universally low confidence in all problem statements. It would therefore be incorrect to interpret these results as identifying which problems are most important for clinicians. The analysis is from the perspective of the audience we intend to teach and educate. Given the established low confidence of this group and immaturity of AI technologies in healthcare, misalignment of importance rating in this survey with the priorities of higher knowledge groups such as AI developers, regulators, academics and clinicians with direct experience of AI integration is expected. The authors do not feel the priorities set by front line clinicians should be considered less important than more specialist groups however, as their understanding of the operational context in which AI systems are to be deployed is essential. Each stakeholder group’s educational priorities for front-line clinicians needs to be evaluated and integrated.

The problem statements are mapped to the DART-Ed Artificial Intelligence and Digital Healthcare Technologies Framework. This involved a systematic review and stakeholder workshops including “early career digital leaders”, “industry representatives”, “educators and subject matter experts”(6). Combined with our own interviews distilling the broader capability statements into 10 contextualized problem statements, there has been considerable input from experts in framing the problems presented to residents and educators. In this respect, “determining an appropriate level of confidence in a decision or result from an AI algorithm” being voted the second highest importance problem shows alignment with the views of experts - as this was identified as a central issue by NHS Digital(19,20).

There is considerable inter-reliance between both problem and capability statements. Our survey results are perhaps best used to understand the learners agenda. By framing AI education for clinicians around problems they identified as most important, content can be designed which engages less popular areas. To give examples:

An appropriate patient explanation about clinical AI tools would require an understanding of mitigating privacy and security risks posed by AI. Patient explanations were rated second least important and training in this isunlikely to be sought out. Liability implications however were rated most important. Highlighting the importance of consent to data sharing in UK GDPR(21) and GMC guidance(2), the necessity for data to be shared with third parties to utilise many AI tools such as AI scribes, and that consent requires notification of this, will highlight the importance of patient explanations by framing it in terms of liability.

Critical appraisal of papers reporting AI interventions was rated least important, whilst determining an appropriate level of confidence in a decision by an AI algorithm was second most important. It is hopefully self-evident that the confidence we should have in an AI algorithm, and crucially the decision to implement such a system, is proportional to the quality of evidence available to support its efficacy. Therefore, teaching core critical appraisal skills for AI systems such as interpreting the quality of evidence for an algorithm(22) can be delivered in the context of interpreting an AI algorithm’s decision.

### Future work

We have reviewed existing frameworks for healthcare AI education, contextualised these for clinicians of the user archetype, and surveyed residents and educators in the region on their priorities and teaching modality preferences. Whilst confidence and importance ratings did not differ between residents and educators, they differed on their preferred teaching approaches. Residents ranked integration into regional teaching highest, whilst educators preferred webinars. A significant proportion of both groups also ranked stand alone courses utilising study leave as first choice - this perhaps identifies a more motivated cohort and demonstrates an appetite for this approach

Our next step will be to design short form teaching sessions delivering basic skills(4) and framed around the liability implications of clinical AI and determining an appropriate level of confidence in a decision made by an AI algorithm. We will initially deliver this in regional teaching to gather feedback. Whilst webinar and e-learning AI educational material is increasingly accessible, there is a clear preference from residents to include this in a live regional teaching format. Crucially, this will facilitate delivery to those unaware or unwilling to access online resources. To enable scaling of training delivery beyond our small regional team, and to share learning from this project, the development of a teaching toolkit is also on the roadmap.

### Issues with survey design

The initial question “are you a resident doctor in the East of England” designed to identify EoE from non-EoE responses was poorly phrased, combining EoE location with resident grade. Consequently, responses were difficult to interpret. In total 20 respondents indicated they were not EoE Residents which does not align with the number of consultants responding demonstrating understandably in-consistent interpretation. As the survey was distributed only within the EoE mailing lists, and the geographical region of respondents was not felt to be of significant importance to the conclusions of the survey we therefore discounted responses to this question from the analysis.

## Conclusion

Clinicians are most likely to be motivated to learn about AI framed around its liability implications and determining appropriate confidence in AI algorithms, as these are perceived as important and confidence is low. Despite being key skills for clinical safety, and low confidence ratings, critical appraisal of AI interventions and patient explanations of AI were rated least important. Residents and Educators have equally low confidence in AI related learning outcomes, highlighting a challenge in education delivery. We will use the results of this survey to iteratively develop regional clinical AI education sessions, initially targeting residents in regional teaching and educators in webinars, and to develop a teaching toolkit to support the scaling of this training effort.

## Supporting information

Appendix 3: CROSS research checklist

Appendix 2: Survey Questionaire

Appendix 1: NHSE DART-Ed Framework Capabiltiy Mapping

## Data Availability

All data produced in the present study are available upon reasonable request to the authors following publication.

