## Appendix 2: Survey Questionaire for "Priorities for AI Education: Clinicians’ Perspectives"

NHS England

### Clinical Artificial Intelligence Learning Needs

Thank you for participating in this NHSE East of England survey to help understand the learning needs of Doctors relating Clinical Artificial Intelligence (AI). We are defining AI broadly, referring to computer systems able to perform human tasks such as image interpretation, decision making and support, speech and language interpretation, communication and robotics.

You will be asked to rate your confidence and rank the importance of ten challenges, as well as your preferred teaching format for this topic.

The information provided by you in this questionnaire will be used for research purposes. It will not be used in a manner which would allow identification of your individual responses.

Contact:
https://innovateeoe.com/


* Required

Are you a resident doctor or educator in the East of England?

- ◯ Yes
- ◯ No

**Challenges:** Please rate your current level of confidence in each of these problems: *

| | 1 (Not at all confident) | 2 | 3 | 4 | 5 (Completely confident) |
|---|---|---|---|---|---|
| I am aware of realistic use cases for AI in healthcare | ◯ | ◯ | ◯ | ◯ | ◯ |
| I can explain to patients how AI works | ◯ | ◯ | ◯ | ◯ | ◯ |
| I am able to write effective prompts for Generative AI and recognise AI generated content | ◯ | ◯ | ◯ | ◯ | ◯ |
| I know common ways that AI can fail | ◯ | ◯ | ◯ | ◯ | ◯ |
| I can identify sources of bias in AI algorithms and potential mitigation measures | ◯ | ◯ | ◯ | ◯ | ◯ |
| I am able to determine an appropriate level of confidence in a decision or result from an AI algorithm | ◯ | ◯ | ◯ | ◯ | ◯ |
| I can critically appraise papers reporting AI interventions in my speciality | ◯ | ◯ | ◯ | ◯ | ◯ |
| I can explain how AI in healthcare is regulated | ◯ | ◯ | ◯ | ◯ | ◯ |
| I know the liability implications of using AI in my practice | ◯ | ◯ | ◯ | ◯ | ◯ |
| I can mitigate security and privacy risks posed by AI | ◯ | ◯ | ◯ | ◯ | ◯ |

Choose up to 3 challenges that you think are most important to address:

Please select at most 3 options.

- ☐ I am aware of realistic use cases for AI in healthcare
- ☐ I can explain to patients how AI works
- ☐ I am able to write effective prompts for Generative AI and recognise AI generated content
- ☐ I know common ways that AI can fail
- ☐ I can identify sources of bias in AI algorithms and potential mitigation measures
- ☐ I am able to determine an appropriate level of confidence in a decision or result from an AI algorithm
- ☐ I can critically appraise papers reporting AI interventions in my speciality
- ☐ I can explain how AI in healthcare is regulated
- ☐ I know the liability implications of using AI in my practice
- ☐ I can mitigate security and privacy risks posed by AI

Rank your preferred teaching formats from most to least preferable.

- During Regional Teaching
- Stand alone course using study leave
- Webinars
- e-Learning modules
- Blogs and social media posts

How could AI improve your work productivity? Provide specific examples or use cases. For example: "Using generative AI to summarise research papers."

What is your training grade?

- ( ) Medical student
- ( ) Foundation Programme
- ( ) ST/CT 1-3
- ( ) ST 4-6
- ( ) ST 7+
- ( ) Consultant
- ( ) Staff Grade
- ( ) Junior Clinical Fellow
- ( ) Senior Clinical Fellow
- ( ) Other

Which speciality training school(s) are you part of?

- [ ] Academic
- [ ] Anaesthetics
- [ ] Dental
- [ ] Emergency Medicine
- [ ] Foundation Programme
- [ ] General Practice
- [ ] Intensive Care Medicine
- [ ] Internal Medicine
- [ ] Obstetrics and Gynaecology
- [ ] Ophthalmology
- [ ] Out of Training
- [ ] Paediatrics
- [ ] Pathology
- [ ] Psychiatry
- [ ] Public Health
- [ ] Radiology
- [ ] Surgery
- [ ] Other

If you have a role in Medical Education, which Tier does this fall into?

- ( ) Tier 1 - Trainees as Educators
- ( ) Tier 2 - Clinical Supervisors
- ( ) Tier 3 - Educational Supervisors
- ( ) Tier 4 - College Tutor or Training Programme Director
- ( ) Tier 5 - Head of School, Associate or Deputy Dean

If you have any ideas, concerns or feedback you would like to communicate please let us know here:

This content is neither created nor endorsed by Microsoft. The data you submit will be sent to the form owner.

Microsoft Forms
