## Appendix 1: NHSE DART-Ed Framework Capabiltiy Mapping for "Priorities for AI Education: Clinicians’ Perspectives"

| Capability | Challenge 1 | Challenge 2 | Challenge 3 |
|---|---|---|---|
| I can confidently explain to patients what AI systems in my specialism/area of practice are doing in a broad sense and can tailor my explanation accordingly with awareness that patients have different levels of digital literacy. | I can explain to patients how AI works | | |
| When applying Artificial Intelligence (AI) processes to direct patient care, I am able to second-check decisions made by automated processes to ensure they meet the required standards of safety and quality and know how and where to report concerns if they fail to meet these standards. | I am able to determine an appropriate level of confidence in a decision or result from an AI algorithm | I know common ways that AI can fail | I know common ways that AI can fail |
| I am aware of the legal, ethical and accountability implications for myself and organisation when following or overruling AI based recommendations in augmented clinical decision making. | I know the liability implications of using AI in my practice | I can identify sources of bias in AI algorithms and potential mitigation measures | I can mitigate security and privacy risks posed by AI |
| I am familiar with the governance requirements (including UK Conformity Assessed (UKCA/UKNI), CE marking) for medical devices including software and AI concerning regulation, quality management, testing and ongoing monitoring. | I can explain how AI in healthcare is regulated | I am aware of realistic use cases for AI in healthcare | |
| I am aware of issues around consent and medical negligence (for example, Bolam, Bolitho, Montgomery v Lanarkshire) and the challenges of applying these to AI and digital technologies. | I know the liability implications of using AI in my practice | I can explain to patients how AI works | I can mitigate security and privacy risks posed by AI |
| I acknowledge the need for multidisciplinary collaboration to ensure the safe and effective use of digital health and Artificial Intelligence (AI) technology. | I can explain how AI in healthcare is regulated | I can mitigate security and privacy risks posed by AI | I can identify sources of bias in AI algorithms and potential mitigation measures |
| I am aware of specialists roles and teams involved in AI and digital healthcare (for example, informaticians, software/data engineers, IT specialists, researchers, specialist clinicians). | I can explain how AI in healthcare is regulated | | |
| I appreciate that responsibly designed and implemented digital technologies and Artificial Intelligence (AI) systems has potential to increase efficiencies, reduce costs and support healthcare staff to deliver better care | I can critically appraise papers reporting AI interventions in my speciality | I am able to write effective prompts for Generative AI and recognise AI generated content | |
| I am able to evaluate the most appropriate technical solution to a problem including AI/digital technology. This includes reviewing financial, technical and logistical considerations. | I can critically appraise papers reporting AI interventions in my speciality | I can mitigate security and privacy risks posed by AI | I can identify sources of bias in AI algorithms and potential mitigation measures |
| I understand that Artificial Intelligence (AI) is an umbrella term used to define digital technologies capable of performing tasks commonly thought to require human intelligence. I am aware AI is common in modern technology and can list uses of AI outside healthcare (for example, voice recognition, recommender systems, self-driving cars, image and video processing) | I can explain to patients how AI works | I am able to write effective prompts for Generative AI and recognise AI generated content | |
| I can provide examples of AI systems used in healthcare and understand their potential benefits and risks (for example, imaging diagnostics and decision support tools) | I am aware of realistic use cases for AI in healthcare | I can critically appraise papers reporting AI interventions in my speciality | |
| I am aware that "machine learning" is a subset of AI and is an umbrella term used to refer to techniques that allow computers to learn from examples/data without being explicitly programmed with step-by-step instructions | I can explain to patients how AI works | | |
| I am aware that all AI applications in healthcare are defined as 'narrow' AI that are trained to perform a particular and specific task | I can explain to patients how AI works | | |
| I can identify the contribution that AI could make to healthcare processes in my area of practice and how it has potential to benefit the organization, workforce and patient | I am aware of realistic use cases for AI in healthcare | | |
| I can articulate the risks and limitations of AI relevant to my professional area and consider them in my use of AI | I know common ways that AI can fail | I can identify sources of bias in AI algorithms and potential mitigation measures | I can mitigate security and privacy risks posed by AI |

| Capability | Challenge 1 | Challenge 2 | Challenge 3 |
|---|---|---|---|
| I can define the sub-fields of AI and machine learning and their key applications (for example, computer vision, audio processing, knowledge representation, natural language processing, expert systems) | I can explain to patients how AI works | | |
| I can describe the main types of bias that could affect AI systems (for example, reporting, selection, group attribution, implicit) | I can identify sources of bias in AI algorithms and potential mitigation measures | | |
| I understand that machine learning algorithms require large quantities of data to learn from, and must be trained and evaluated using independent sub-sets of the available data | I can critically appraise papers reporting AI interventions in my speciality | I can identify sources of bias in AI algorithms and potential mitigation measures | |
| I am aware of some of the common uses for Natural Language Processing (NLP) methods and text mining within and outside of healthcare (for example, chat bots, speech/virtual assistants, dictation of clinical notes, processing electronic patient records) | I am aware of realistic use cases for AI in healthcare | I can explain to patients how AI works | |
| I am aware of the use of virtual assistants (for example, Amazon Alexa, Google Assistant) in healthcare to improve accessibility for patients (for example, patients with disabilities) to access health information and can recommend their use to patients where appropriate | I can explain to patients how AI works | | |
| I am able to use Artificial Intelligence (AI) systems confidently to assist me to improve task efficiency while maintaining quality and safety | I am able to write effective prompts for Generative AI and recognise AI generated content | I can mitigate security and privacy risks posed by AI | I know the liability implications of using AI in my practice |
| I know how to respond if an AI system fails or is inaccessible and can initiate an alternative process to maintain effective service provision | I know common ways that AI can fail | I am able to determine an appropriate level of confidence in a decision or result from an AI algorithm | |
| I understand the importance of sharing learning following failures of an AI system, to improve systems and practice | I know common ways that AI can fail | I am able to determine an appropriate level of confidence in a decision or result from an AI algorithm | I am aware of realistic use cases for AI in healthcare |
| I am aware of the limitations of AI systems and how to respond when AI derived information contradicts my clinical/professional intuition. I retain a 'critical eye' and am aware of how AI may influence my decision making | I am able to determine an appropriate level of confidence in a decision or result from an AI algorithm | I know common ways that AI can fail | |
| I actively maintain my clinical knowledge and skills to ensure that my clinical performance is not adversely affected by de-skilling resulting from using AI | I know the liability implications of using AI in my practice | | |
| I can set thresholds for monitoring patients using AI enabled decision support systems for chronic health conditions to generate alerts to initiate appropriate action (for example, call patients in for review, alter treatment) | I am able to determine an appropriate level of confidence in a decision or result from an AI algorithm | I can identify sources of bias in AI algorithms and potential mitigation measures | |
| When evaluating an AI system for use in my professional workflow, I can compare its performance against the expected standards in my professional area of practice | I can critically appraise papers reporting AI interventions in my speciality | | |
| I am aware of the use of robotic technology for healthcare and can cite examples of the use of robots for health, medical and social care (for example, social companion robots, surgical robots, room disinfectant robots, nanotechnology) | I can explain to patients how AI works | | |
| I am aware of the uses of telepresence robots to carry out basic procedures (for example, temperature monitoring) in highly infectious patients and for use with elderly to support independence or for remote care and assessment | I can explain to patients how AI works | | |
| I know where and how to access training (local or manufacturer provided) on robotic technology that I am expected to use | I know common ways that AI can fail | I can mitigate security and privacy risks posed by AI | |
| I acknowledge the need for multidisciplinary collaboration to ensure the safe and effective use of digital health and Artificial Intelligence (AI) technology. | I am aware of realistic use cases for AI in healthcare | I can explain how AI in healthcare is regulated | |
